# Optical brain pulse monitoring of microvascular blood flow during endovascular treatment for acute ischemic stroke

**DOI:** 10.1101/2025.02.04.25320869

**Authors:** Sigrid Petautschnig, Elliot J Teo, Lauren Sanders, Ashu Jhamb, Julian Maingard, Finula I Isik, Jean Lee, Barry Dixon

**Author notes:** Corresponding author Correspondence: Dr Barry Dixon L18, 1 Nicholson St, CYBAN, East Melbourne 3002 Co-Corresponding author: Dr Sigrid Petautschnig.

## Abstract

**Background:** Endovascular treatment (EVT) of large vessel occlusion (LVO) strokes improves patient outcomes. However, significant challenges remain including detection of microvascular no reflow phenomena, emboli to new territories and improving neuro-prognostication. Real time monitoring of the brain microcirculation could assist in addressing these challenges. This first-in-human cohort study evaluated Optical Brain Pulse Monitoring (OBPM) of microvascular blood flow during EVT.

**Methods:** OBPM is a non-invasive device using red and infrared light to capture brain pulse waveforms, reflecting the relative arteriole and venous pressure levels driving microvascular blood flow. OBPM brain pulse classes – Arterial, Hybrid, Venous I, Venous II and Monotonous – represent a continuum of blood flow states from normal (Arterial) to critically low blood flow (Monotonous). OBPM sensors were positioned bilaterally over the middle cerebral artery territories of LVO stroke patients undergoing EVT. Data on patient demographics, imaging and clinical outcomes were collected.

**Results:** Eleven patients (mean age 71, NIHSS 13) were enrolled. The most common brain pulse class at presentation was the Venous I (64%). The brain pulse class at presentation was associated with the hypoperfused tissue volume (p = 0.005). The brain pulse class following EVT was associated with long-term patient outcomes, including length of hospital stay (p = 0.04), the modified Rankin Score (p = 0.06) and hospital death (p = 0.02). In one patient, OBPM detected an embolization to a new territory that occurred during internal carotid artery stenting.

**Conclusion:** The OBPM brain pulse waveforms demonstrated venous circulation features in patients presenting with LVO stroke. These waveforms were also associated with clinical outcomes, such as stroke size at presentation, hospital LOS and mortality. Monitoring of the brain pulse during EVT could improve intra-procedural evaluation of microvascular no reflow, detection of complications and neuro-prognostication. OBPM also has potential as a simple method for earlier LVO stroke detection.

**Clinical trial registration:** URL:https://www.anzctr.org.au/Trial/Registration/TrialReview.aspx?id=384769&isReview=true; Unique identifier: ACTRN12622001320741

## Introduction

Endovascular treatment (EVT), particularly mechanical thrombectomy, has emerged as a cornerstone in the management of acute ischemic stroke due to large vessel occlusion (LVO).^1^ EVT is a minimally invasive procedure that involves navigating a catheter through the vascular system to the site of the occlusion, enabling the mechanical removal of the clot and restoring cerebral blood flow. The efficacy of EVT has been demonstrated in numerous clinical trials, with outcomes showing reduced mortality and improved patient functional recovery compared to thrombolytic management alone.^2^

The technical success of EVT is traditionally measured by the achievement of reperfusion, evaluated using the Modified Treatment in Cerebral Ischemia (mTICI) Scale which is qualitatively estimated by the interventionalist at the end of EVT from the final angiography image. However, there remains a gap between successful arterial reperfusion as assessed by the mTICI grade and good patient outcomes, as almost half of these patients do not experience favorable outcomes despite successful thrombectomy.^3–5^ Limitations in mTICI assessment of microvasculature reperfusion may be an important factor in this inconsistency.

Microvascular no reflow phenomena (NRP) is common following EVT.^5,6^ Computed tomography (CT) and magnetic resonance imaging (MRI) perfusion studies found NRP occurred in up to 50% of patients despite achieving a mTICI score of 3, indicating 100% arterial reperfusion.^6–8^ The presence of NRP is associated with greater infarct growth, hemorrhagic transformation and increased risk of being dependent or dead at 90 days.^6,9^ Proposed mechanisms of NRP include ischemic reperfusion injury with microvascular thrombosis, neutrophil margination in post capillary venules and pericyte mediated capillary constrictions.^10,11^ New embolization of clot fragments during EVT may also contribute to NRP.^10^ This occurs in up to 33% of patients and is difficult to identify intra-procedurally.^12^

Optical Brain Pulse Monitoring (OBPM), developed by Cyban Pty Ltd (Melbourne, Australia), represents a promising novel approach to monitoring microvascular cerebral blood flow responses to EVT compared with traditional digital subtraction angiography (DSA) and mTICI score. OBPM aims to provide continuous monitoring of cerebral blood flow during stroke treatment. Alternative monitoring options, such as cerebral oximetry and EEG, have not demonstrated clinical efficacy to date.^13^

OBPM uses red and infrared light to capture brain pulse waveform classes, whose morphology reflect the relative arteriole to venous pressure levels that drive microvascular blood flow.^14^ Previous preclinical and clinical work using the OBPM has identified brain pulse classes including Arterial, Hybrid, Venous I, Venous II and Monotonous, ranging from normal (Arterial) to critically low blood flow (Monotonous) – that represent a continuum of blood flow states from normal (Arterial) to critically low blood flow (Monotonous, Figure 1A-E).^14–18^

**Figure 1.**
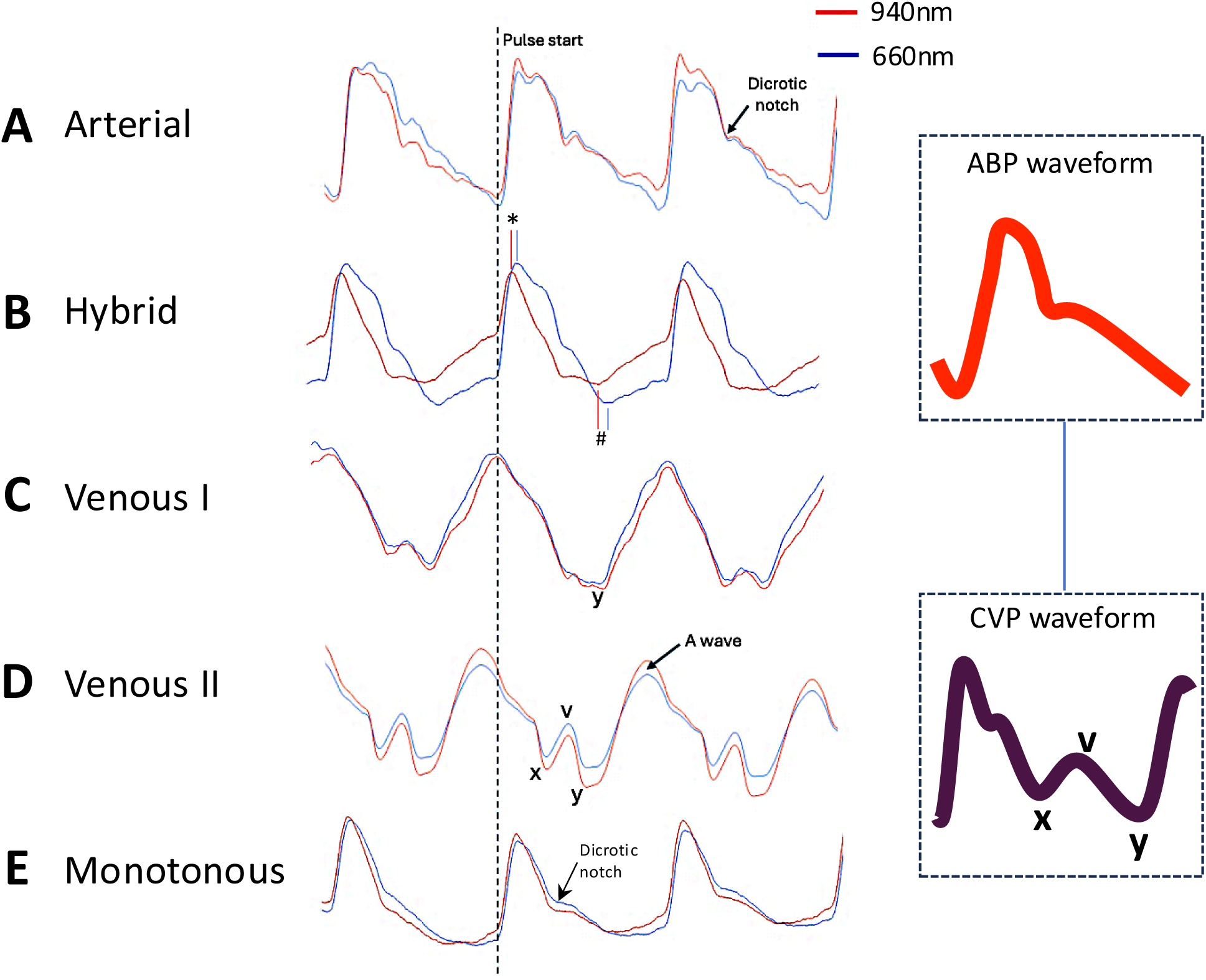
Summary of brain pulse waveform classes observed in acute ischemic stroke by OBPM. A) An Arterial brain pulse which has a similar shape to a normal arterial pressure waveform. B) The Hybrid brain pulse is characterized by distinct shapes for the 940 nm and 660 nm wavelengths, due to their different responses to low blood oxygen levels. The pulse peaks * and troughs # of 660 nm are delayed relative to 940 nm. C) The Venous I brain pulse may have somewhat undifferentiated features, consistent with similar arteriole and central venous pressure levels throughout the cardiac cycle. The pulse may demonstrate more obvious CVP features in the diastolic phase including V and Y waves. D) The Venous II brain pulse has clearer CVP features such as the A, C, V, X and Y waves. The Venous II pulse is consistent with the venous pressure exceeding arteriole pressure throughout the cardiac cycle. E) Monotonous brain pulse has absence of variation in shape between pulses and a low dicrotic notch. Red brain pulse is 940 nm and blue brain pulse 660 nm. *Abbreviations: OBPM, Optical Brain Pulse Monitor; CVP, central venous pressure*.

The OBPM’s optical signal reflects pulsatile blood volume changes in the pial venules that lie on the cortical surface within the sub-arachnoid space (Figure 2A). The pial venules relative blood volume is 4-fold higher than in the capillary beds of the cortex.^19^ The pial venules act as a blood reservoir.^20,21^ Thus, a very large venous blood volume lies on the surface of the cortex, providing an optical signal. Functional MRI studies have demonstrated that the pial venous blood is dynamic in response to physiological changes in blood flow and dilate acutely following ischemic stroke.^20,22,23^

**Figure 2.**
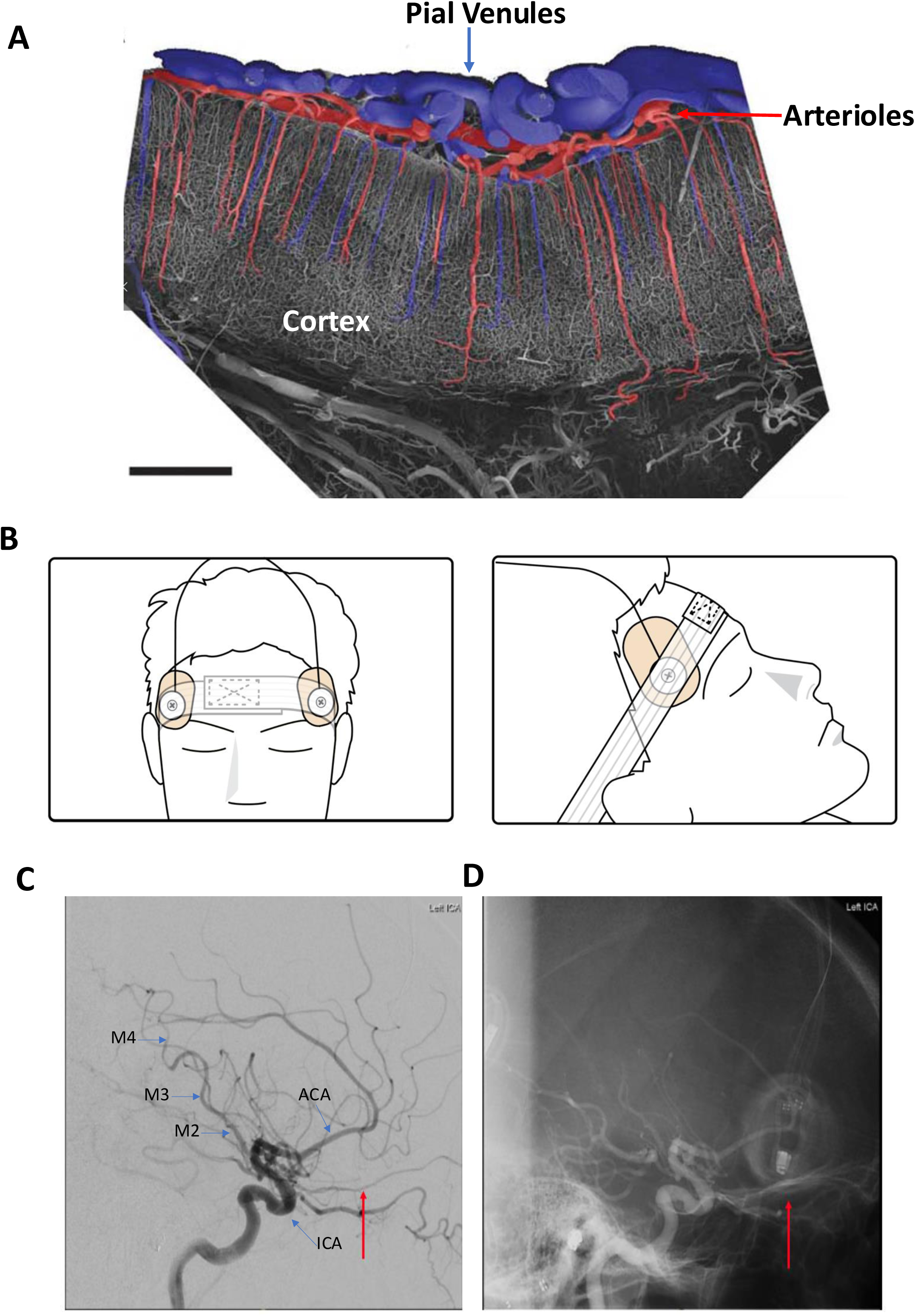
OBPM sensor placement. (A) Scanning electron micrograph of a vascular corrosion cast from the monkey visual cortex (primary visual cortex). Arterioles are red and the pial venules are blue. Bar =1LJmm. Image adapted from Hirsch et al. 2012 with permission from SAGE publishing. (B) OBPM sensor and adhesive placement over the anterior temporal region. (C) Representative positive and (D) negative DSA contrast images. Red arrows indicate the OBPM sensors position. *Abbreviations: OBPM, Optical Brain Pulse Monitor; ICA, internal carotid artery; MCA, middle cerebral artery; DSA, digital subtraction angiography*.

OBPM has been extensively assessed in critically ill patients with acute brain injury.^16,24–26^ A submitted study in an ovine ischemic stroke model provided proof of concept that middle cerebral artery (MCA) stroke was associated with venous circulation changes in the OBPM brain pulse waveform.^18^

The primary aim of this first-in-human stroke study was to assess the proportion of patients in which an OBPM brain pulse was obtained, assessed by the morphology of the pulse waveform classes, as observed in previous studies.^14,17,18^ Additional exploratory aims were to assess the association between the brain pulse waveform classes with stroke tissue volumes at presentation and clinical outcomes following EVT.

## Methods

The study was conducted at St Vincent’s Hospital Melbourne, Australia. Patients were enrolled during office hours between July 2022 and July 2024. St Vincent’s Hospital (Melbourne, Australia) Human Research Ethics Committee (HREC) granted ethics committee approval (HREC 231/22 and HREC 160/20). Informed consent was obtained from the participant or, in cases where participants lacked the capacity to consider participation at the time of eligibility, the participant’s medical treatment decision maker. In the event the participants medical treatment maker could not be contacted prior to thrombectomy, deferred consent was obtained. All medical protocols in this study adhered to the Declaration of Helsinki and the International conference on Harmonization guidelines for Good Clinical Practice (ICH-GCP).

### Participants

Patients presenting within 24 hours of last known well undergoing EVT for LVOs in the internal carotid artery (ICA) and/or M1/M2 segments of the MCA were included in this study. Patients were excluded if they had a pre-existing major brain injury or hemorrhage, as these could affect the baseline OBPM measurements.

### Data collection

All patients underwent CT brain, CT angiography and CT perfusion imaging and a stroke was clinically diagnosed by the treating clinicians. The hypoperfused tissue volume (Tmax > 6 seconds), core volume (<30% normal cerebral blood flow) and the penumbra volumes were assessed by CT perfusion with automated postprocessing by the RAPID software (iSchemaView, Menlo Park, CA).

Upon patient arrival to the DSA suite, the OBPM sensors were placed bilaterally on the anterior temporal region of the scalp, over the MCA territories (Figure 2B-D). The sensors were maintained in position with a head band and adhesive tab. All OBPM sessions began after intubation (if undertaken) and prior to the start of EVT (defined as groin puncture) and ended shortly before patient departure from the DSA suite.

The EVT procedures utilized stent retriever devices, thrombus aspiration, angioplasty and stenting, or a combination of techniques. The choice of the EVT thrombectomy device and treatment technique was at the discretion of the neuro-interventional radiologist.

Demographic, procedural (including mTICI scores), complication, anesthetic and outcome data were collected at the time of EVT or from the hospital electronic patient record. Significant clinical timepoints (including clot aspiration, stent deployment, balloon inflation) throughout the procedure were recorded in real time by a member of the research team (EJT). Patient outcomes included the modified Rankin scale (mRS) at 90 days after EVT, length of hospital stay (LOS), and hospital survival. For patients that died in hospital the LOS was adjusted to maximum of the surviving cohort (55 days) for statistical analyses.

### Optical brain pulse monitor

The OBPM is comprised of a roll-stand with an enclosure containing the Graphical User Interface (GUI) (a Tablet PC), a power supply for all components. The monitor’s LED and PD are controlled and processed by an Integrated Analog Front End circuit board, which digitizes the received signal from each sensor and sends the data stream to the Tablet PC. The PC receives the sensor data from the Processing Unit, and presents the data on a display, along with patient identifier data to the operator, via a custom software application. Data from the OBPM was captured at a rate of 500 Hz and stored for subsequent analysis. This analysis was conducted using Python v3.10, SciPy v1.10. The clean data were filtered to highlight specific frequency components relevant to physiological mechanisms for quantitative analyses. Synchronous recordings were made with the routine intensive care monitoring outputs, such as blood pressure, central venous pressure, ECG, end tidal CO_2_ and heart rate. The physiological data was exported from the Philips IntelliVue system using ICM+ (Cambridge Enterprise, Cambridge, UK).

The brain pulse waveform class of the OBPM outputs were qualitatively assessed over the hemispheres ipsilateral and contralateral to the stroke. Waveforms were reviewed and classified based on their morphology compared with previously defined waveform classes.^14,16,25^ These recordings were examined by three expert observers (EJT, SP, BD). The observers, blinded to the patients CT finding and the procedural timepoint, classified the predominant waveform type into five distinct classes: 1) Arterial, 2) Hybrid, 3) Venous 1, 4) Venous II and 5) Monotonous.

The primary outcome was the proportion of patients with LVO stroke in which a brain pulse could be detected. This outcome was chosen based on the available resources that constrained the sample size to 10 to 15 patients.

### Exploratory analyses

To assess brain pulse class responses during EVT analysis was undertaken at 4 points of the brain monitoring session. 1) at the beginning of the monitoring episode, 2) just prior to clot retrieval, 3) shortly following clot retrieval, and 4) at the end of the monitoring episode. The responses were categorized into 4 categories. 1) Brain pulse class improved, 2) Brain pulse class worsened during EVT but reverted to the class at start, 3) Brain pulse class did not change throughout EVT, 4) Brain pulse class worsened from the start of EVT to the end.

The association of the brain pulse class at presentation with the stroke tissue volumes was assessed. The brain pulse class at the end of EVT and the mTICI associations with long term patient outcomes were also assessed.

As one patient had two EVT procedures, long-term patient outcomes were only related to brain pulse waveforms taken from their second EVT, given the final revascularization status post-treatment and is likely more relevant to long-term outcomes than prior treatments. This approach assumes that the final successful EVT has a dominant effect on outcomes.

### Statistical analysis

Interrater reliability in brain pulse waveform class was analyzed using the “irr” package v0.84.1 in R studio using the Fleiss κ-statistic. Other statistical analyses were undertaken using GraphPad Prism Version 10.4.1. Statistical significance was assessed by Welch’s t-test, Kruskal-Wallis one-way ANOVA or Fischer’s exact test. The effect of time point on the brain pulse class comparing the ipsilateral and contralateral hemispheres groups was assessed with repeated measures analysis, where an ordinal scale was used to represent the brain pulse class. The exploratory analyses did not correct for multiple comparisons.

## Results

### Patient characteristics

Eleven adult patients with acute LVO strokes were studied. The mean age was 70.6 years (SD ± 12.1 years), 6 were female and the mean baseline NIHSS was 13 (Table 1), the mean last seen well to the start of EVT time was 5 hours and 6 minutes (range: 43 mins – 18 hours). Hospital LOS mean was 18 days and ranged from 3 – 55 days, and hospital survival was 73% (Supp. Table 1).

**Table 1.**
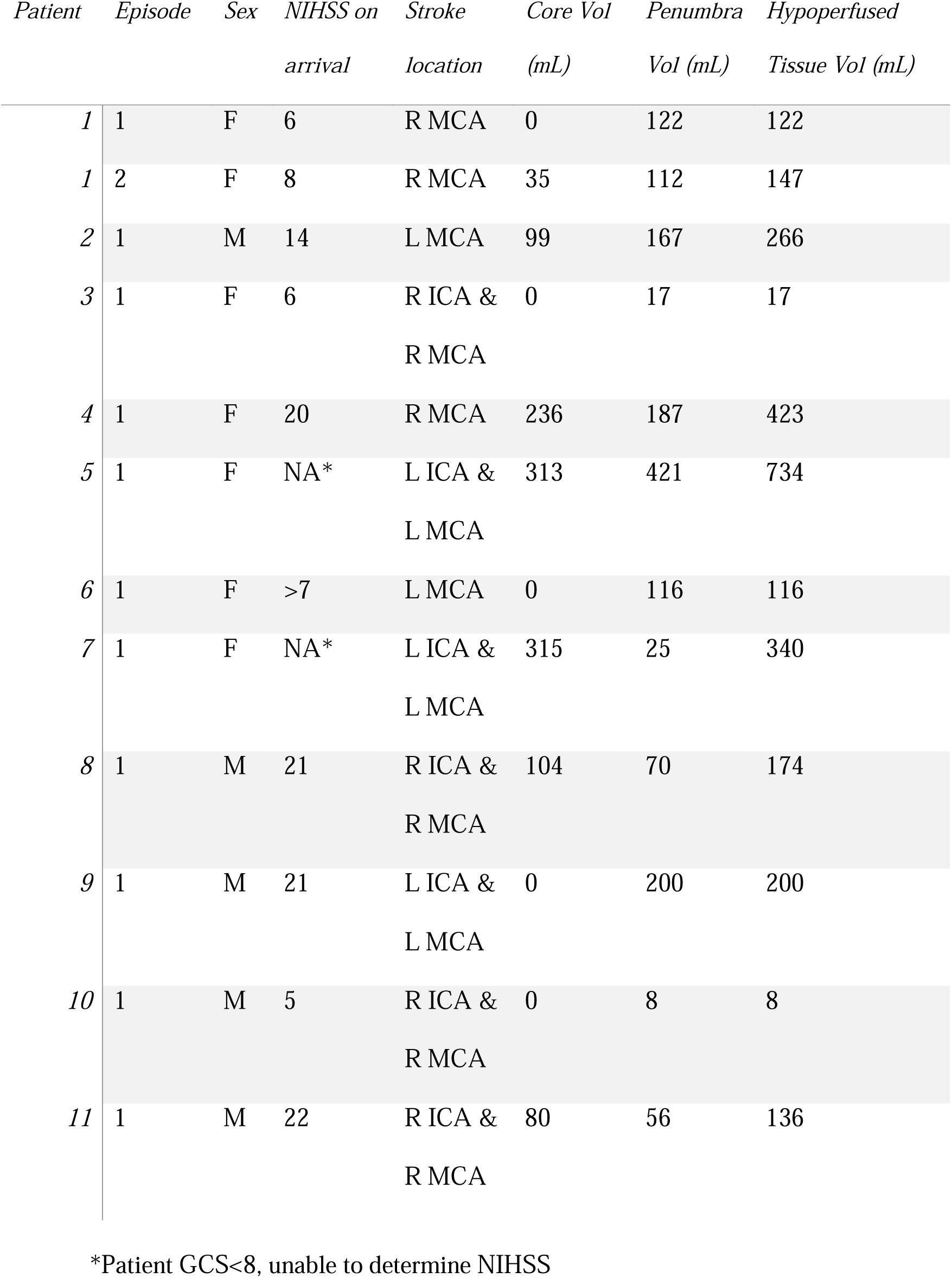
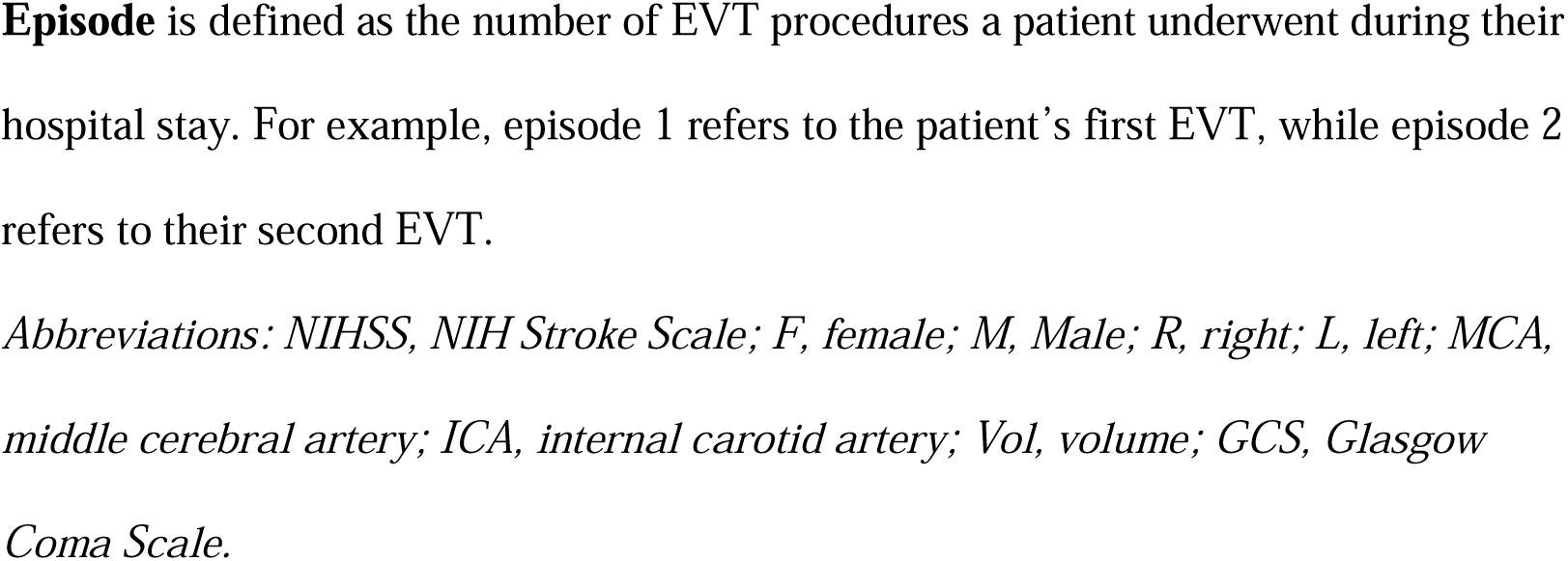
Demographics and infarct characteristics of patients.

| *Patient* | *Episode* | *Sex* | *NIHSS on*<br>*arrival* | *Stroke*<br>*location* | *Core Vol*<br>*(mL)* | *Penumbra*<br>*Vol (mL)* | *Hypoperfused*<br>*Tissue Vol (mL)* |
| --- | --- | --- | --- | --- | --- | --- | --- |
| *1* | 1 | F | 6 | R MCA | 0 | 122 | 122 |
| *1* | 2 | F | 8 | R MCA | 35 | 112 | 147 |
| *2* | 1 | M | 14 | L MCA | 99 | 167 | 266 |
| *3* | 1 | F | 6 | R ICA &<br>R MCA | 0 | 17 | 17 |
| *4* | 1 | F | 20 | R MCA | 236 | 187 | 423 |
| *5* | 1 | F | NA* | L ICA &<br>L MCA | 313 | 421 | 734 |
| *6* | 1 | F | >7 | L MCA | 0 | 116 | 116 |
| *7* | 1 | F | NA* | L ICA &<br>L MCA | 315 | 25 | 340 |
| *8* | 1 | M | 21 | R ICA &<br>R MCA | 104 | 70 | 174 |
| *9* | 1 | M | 21 | L ICA &<br>L MCA | 0 | 200 | 200 |
| *10* | 1 | M | 5 | R ICA &<br>R MCA | 0 | 8 | 8 |
| *11* | 1 | M | 22 | R ICA &<br>R MCA | 80 | 56 | 136 |

\*Patient GCS<8, unable to determine NIHSS
**Episode** is defined as the number of EVT procedures a patient underwent during their hospital stay. For example, episode 1 refers to the patient's first EVT, while episode 2 refers to their second EVT.
*Abbreviations: NIHSS, NIH Stroke Scale; F, female; M, Male; R, right; L, left; MCA,* *middle cerebral artery; ICA, internal carotid artery; Vol, volume; GCS, Glasgow* *Coma Scale.*

OBPM episodes were recorded in all patients across 12 EVT procedures (Patient 1 had two strokes during the hospital admission). The OBPM monitoring session mean time was 68 minutes and ranged from 26 to 117 minutes. The EVT procedures involved clot aspiration, stenting and balloon angioplasty. Localization of clot was reported in the MCA in five instances, and in both the MCA and ICA in the remaining seven. The hypoperfused tissue volume ranged from 8 – 734 mL (Table 1). Two patients also underwent thrombolysis prior to EVT, and ten of the 11 patients were mechanically ventilated during the procedure (Supp. Table 2).

**Table 2.** Ipsilateral brain pulse waveform class over the course of EVT.

| *Patient* | *Episode* | *Start of EVT* | *During clot*<br>*retrieval* | *Post-clot*<br>*retrieval* | *End of EVT* | *Change in*<br>*class* |
| --- | --- | --- | --- | --- | --- | --- |
| **1** | 1 | Venous I | Venous I | Venous I | Venous I | No change |
| **1** | 2 | Venous I | Arterial | Arterial | Hybrid | Improved |
| **2** | 1 | Venous I | Venous II | Venous II | Venous I | Reversion |
| **3** | 1 | Arterial | Hybrid | Arterial | Arterial | Reversion |
| **4** | 1 | Venous I | Venous I | Venous I | Venous I | No change |
| **5** | 1 | Monotonous | Monotonous | Monotonous | Monotonous | No change |
| **6** | 1 | Arterial | Arterial | Arterial | Monotonous | Worsened |
| **7** | 1 | Venous I | Arterial | Venous I | Venous II | Worsened |
| **8** | 1 | Venous I | Venous I | Venous I | Venous I | No change |
| **9** | 1 | Venous I | Venous I | Venous I | Venous I | No change |
| **10** | 1 | Arterial | Venous I | Venous II | Arterial | Reversion |
| **11** | 1 | Hybrid | Hybrid | Venous I | Venous I | Worsened |

**Episode** is defined as the number of EVT procedures a patient underwent during their hospital stay. For example, episode 1 refers to the patient's first EVT, while episode 2 refers to their second EVT.
*Abbreviations: EVT, Endovascular treatment*

### Brain pulse class at presentation

Brain pulse waveforms were observed in all patients using OBPM. Brain pulse classifications between expert observers demonstrated moderate agreement (*κ* = 0.6, *p <* 0.0001). Abnormal brain pulse classes were present on both the ipsilateral and contralateral sides of the stroke. No significant differences in brain pulse classes were found between hemispheres at presentation or during EVT (p = 0.9)

The most common ipsilateral brain pulse class at presentation was Venous I (64%). The brain pulse class at the start of EVT was strongly associated with the hypoperfused tissue volume (Figure 3, p = 0.005). The association was not significant for penumbra (p = 0.08) or the core volume (p = 0.11). The brain pulse class contralateral to the stroke was associated with the penumbra volume (Supp. Figure 1A, p = 0.02), but was not associated with the with the hypoperfused (p = 0.10) or core volumes (p = 0.46).

**Figure 3.**
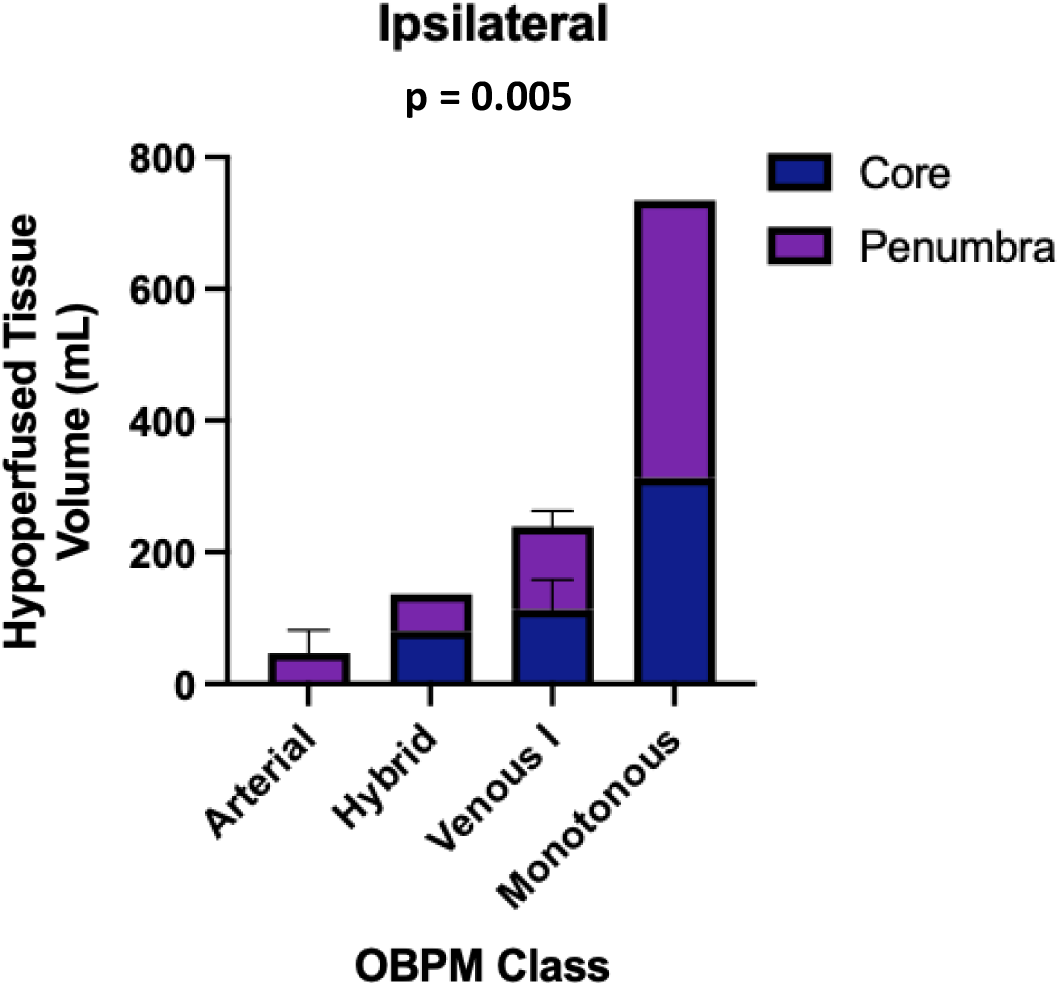
Hypoperfused Stroke Volume (Tmax > 6 seconds) association with the OBPM Class at presentation. Hypoperfused stroke volume was associated with ipsilateral brain pulse waveform class (p = 0.005, Kruskal–Wallis test) at presentation. The components of the hypoperfused stroke volume, Core (<30% normal cerebral blood flow) and penumbra are demonstrated. *Abbreviations: OBPM, Optical Brain Pulse Monitor*.

### OBPM class responses to EVT

On the ipsilateral side (Table 2), over the course of EVT, the brain pulse class improved in one patient, three patients demonstrated a deterioration followed by a reversion back to the brain pulse class at the start of EVT, four patients displayed no change in the brain pulse class and three patients displayed worsening of class from the beginning to the end of EVT. Patients that demonstrated either an improvement in class or a brief deterioration followed by reversion had a shorter hospital LOS compared to patients in which the class did not change or worsened during EVT (p = 0.02, Supp. Figure 1B).

On the contralateral side (Supp. Table 3), the class deteriorated during EVT but reverted to the brain pulse at start in four patients, five patients displayed no change, and two patients displayed worsened class. Like the ipsilateral side, patients that demonstrated a brief deterioration followed by reversion in brain pulse class on the contralateral side had a shorter hospital LOS compared to patients in which the contralateral class did not change or worsened during EVT (p = 0.01, Supp. Figure 1C).

### mTICI responses to EVT

Ten of 11 patients demonstrated successful reperfusion achieving a mTICI score of 2B or greater (indicating > 50% arterial reperfusion). One patient had a poor reperfusion score achieving a mTIC1 of 1.

### Long term patient outcomes

Brain pulse classes on the ipsilateral side at the end of EVT were associated with hospital LOS (p = 0.04, Figure 4A). A potentially clinically relevant effect was present for the 90-day mRS score, but did not reach statistical significance (p = 0.06, Figure 4B). All patients with Arterial, Hybrid, or Venous I survived their hospital stay, while all patients with Venous II or Monotonous brain pulse classes did not (p = 0.02, Figure 4C).

**Figure 4.**
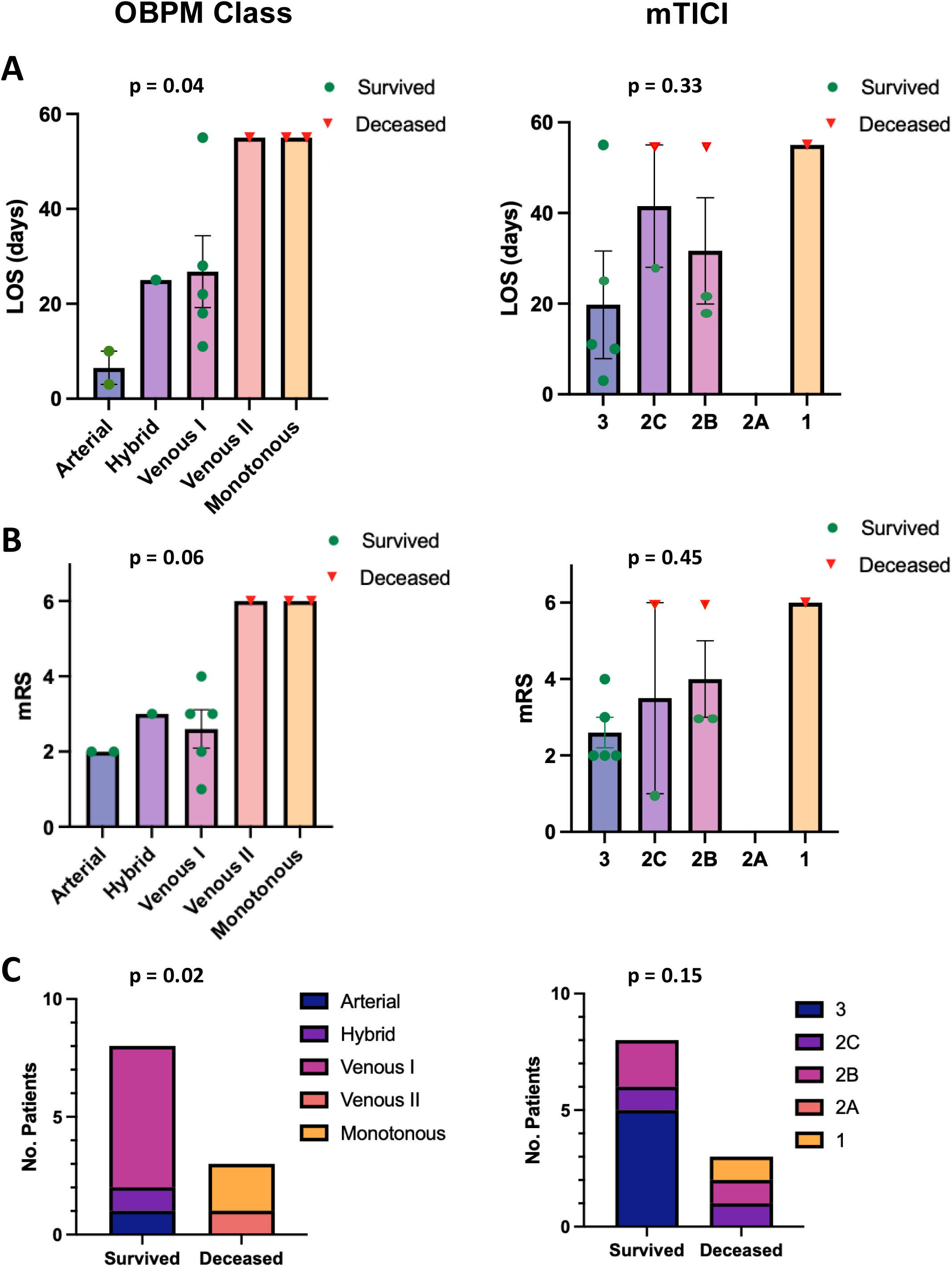
Long-term Patient Outcomes associations with Ipsilateral Optical Brain Pulse Class at end of EVT (left panel) and mTICI Score (right panel). (A) Adjusted hospital LOS (OBPM p = 0.04, mTICI p = 0.33), (B) mRS at 90 days (OBPM p = 0.06, mTICI p = 0.45), (C) In-hospital mortality (OBPM p = 0.02, mTICI p = 0.15). OBPM demonstrated statistical associations with long term patient outcomes but mTICI did not, assessed using (A-B) Kruskal–Wallis test and (C) Fischer’s exact test. *Abbreviations: mTICI, modified treatment in cerebral infarction; LOS, length of stay; EVT, endovascular therapy; mRS, modified Rankin Scale*.

On the contralateral side, only the 90-day mRS score demonstrated an association with the brain pulse class at the end of EVT (p = 0.04, Supp. Figure 2A). There were no associations with hospital LOS or survival (Supp. Figures 2B and 2C).

There were no significant associations between the mTICI score with hospital LOS (p = 0.36, Figure 4A), the 90-day mRS (p = 0.45, Figure 4B) or hospital death (p = 0.15, Figure 4C).

### OBPM and mTICI responses to EVT in cases of hospital death

Three patients in this study (Patient 5, Patient 6 and Patient 7) did not survive their hospital stay. Patient 7, a post-cardiac surgery patient, experienced a sudden drop in Glasgow coma scale (GCS) in the intensive care unit. CT imaging demonstrated a LVO stroke with a large hypoperfused tissue volume (340 mL, Figure 5A). The patient underwent EVT achieving a mTICI grade of 2C, indicating near-complete arterial reperfusion (Figure 5B and 5C). The patient however had no change in the brain pulse class throughout EVT with persistent bilateral Venous II brain pulses, suggesting ongoing poor microvascular blood flow (Figure 5B and 5C). Two days later, CT imaging revealed a large MCA infarction, accompanied by edema with midline shift. The patient transitioned to end-of-life care (Figure 5D).

**Figure 5:**
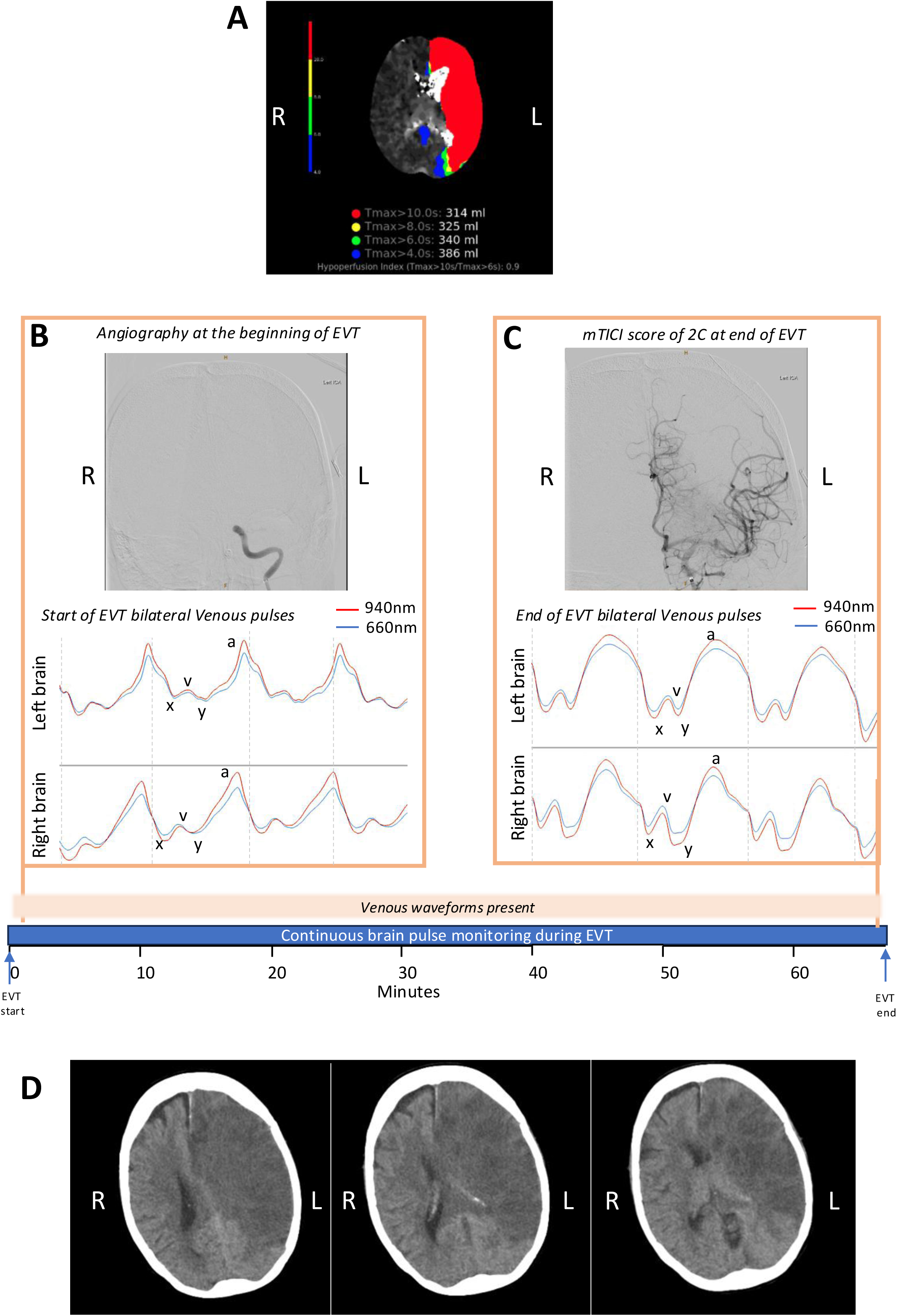
Distinct responses of DSA and brain pulse waveforms to EVT in patient with poor neurological outcome. (A) Rapid CT perfusion imaging demonstrates large left hypoperfused volume at presentation. (B) At the start of EVT, angiography demonstrates occlusion of left ICA and OBPM demonstrates bilateral Venous II brain pulses. (C) At the end of EVT, DSA demonstrates good arterial response with mTICI 2C achieved but OBPM did not respond. Faint grey dashed lines represent the start of each brain pulse. The *a* and *v waves,* and *x* and *y* descents are annotated for a single left and right Venous II pulse. (D) CT images 2 days post-EVT, showing established left MCA infarct with edema and midline shift, the patient did not survive. Red brain pulse is 940 nm and blue brain pulse 660 nm*. Abbreviations: DSA, digital subtraction angiography; EVT, endovascular therapy; mTICI, modified treatment in cerebral infarction; CT, computed tomography; ICA; internal cerebral artery; MCA, middle cerebral artery; L, left; R, right*.

Patient 5 presented with an extremely large hypoperfused tissue volume of 734 ml involving both hemispheres. The initial CT scan demonstrated an acute left ICA occlusion and an associated chronic right ICA occlusion (Supp. Figure 3A). Following EVT the patient achieved a successful mTICI of 2C. The patient however had no change in the brain pulse class with a persistent left sided Monotonous brain pulse, suggesting absent microvascular blood flow (Supp. Figure 3B and 3C). The following day CT demonstrated an established infarct of the entire left hemisphere with midline shift, the patient transitioned to end of life care (Supp. Figure 3D).

Patient 6, with a history of vascular dementia, developed a Monotonous brain pulse during EVT and subsequently died in hospital. On presentation the CT scan demonstrated a left M1 occlusion with extensive chronic white matter ischemia of both hemispheres and the hypoperfused volume was 116 ml. During EVT the occluded vessel could not be opened despite multiple passes due to severe underlying intracranial atherosclerotic disease. A deployed stent also did not open, and the final result was a mTICI of 1. CT imaging at 24 hours demonstrated an established large L MCA infarct of the temporal lobe. In view of her age and co-morbidities the patient transitioned to end of life care.

### Emboli to new territory

An embolus to new territory occurred in Patient 10 during ICA stenting. The patient had undergone uneventful aspiration of a M2 thrombus, subsequently during stenting of the ICA the OBPM waveform class deteriorated acutely from Arterial to Venous II (Figure 6A). Balloon inflation during stenting resulted in a brief period (12 seconds) of bradycardia and a brief 46 second drop in mean arterial blood pressure to 53 mmHg (Figures 6B). Nine minutes and 30 seconds later, a clot in the right M1 territory was identified on a routine DSA run. The Venous II brain pulse improved back to Arterial following aspiration of the clot, a further 12 minutes later (Figure 6A).

**Figure 6:**
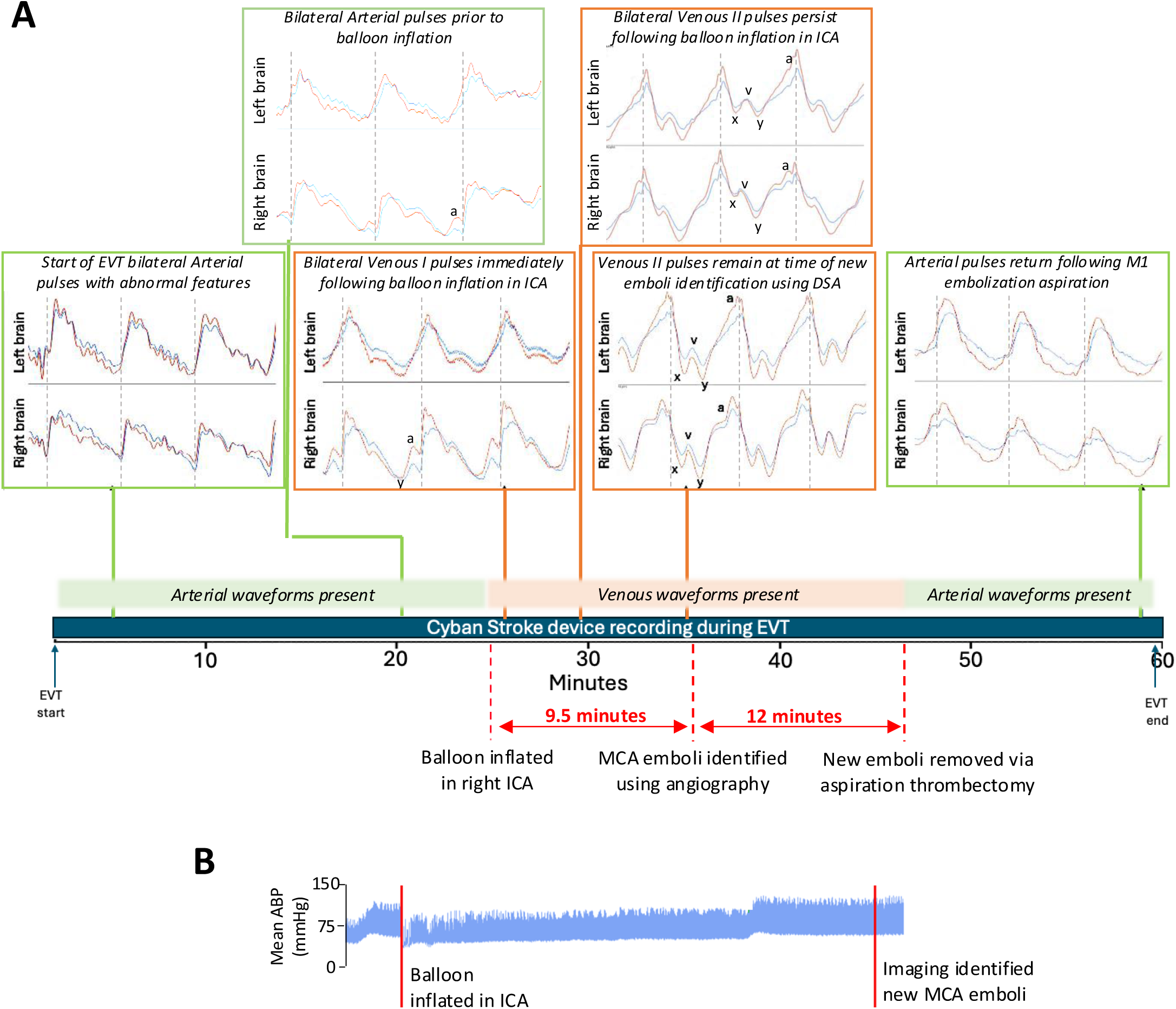
OPBM brain pulse classes change from Arterial to Venous II immediately following new embolization of the right M1 during ICA stenting. (A) In a patient with a right-side small volume 8 mL stroke, stenting of the right ICA stenosis triggered an acute change in the brain pulse class from Arterial to Venous II. Nine minutes and 30 seconds following the change in brain pulse class, a new embolus in the MCA M1 territory was identified during a routine DSA run. Bilateral Arterial brain pulses returned after the M1 embolus was successfully aspirated 12 minutes later. (B) Arterial blood pressure changes associated with ICA stenting. Faint grey dashed lines mark the OPBM brain pulse starts. The *a* and *v waves,* and *x* and *y* descents are annotated. Red brain pulse is 940 nm and blue brain pulse 660 nm*. Abbreviations: ICA, internal carotid artery; MCA, middle cerebral artery; EVT, endovascular therapy*.

## Discussion

This first-in-human study found the OBPM brain pulse waveform had venous circulation features in acute LVO stroke. The waveforms were associated with other important clinical outcomes, such as stroke size at presentation, hospital LOS and mortality. Monitoring of the brain pulse during EVT could improve intra-procedural evaluation of microvascular no reflow, detection of complications and neuro-prognostication. OBPM may also have potential as a simple point of care method for LVO stroke detection in resource limited settings.

### The pathophysiology of the OBPM brain pulse classes

We believe the major cerebrovascular factor influencing the OBPM waveform shape, and therefore OBPM class, is the relative arteriole to venous pressure levels acting across the pial venule microvascular beds of the brain that drive microvascular blood flow.

The *Arterial brain pulse* is typically seen in normal brains with adequate cerebral blood flow and has an arterial pressure pulse shape (Figure 1A).^14^ The shape of the Arterial waveform likely reflects the blood pressure driven increase in the cortical blood volume, which in turn compresses and empties the low pressure pial venules that sit adjacent to the cortical surface (Figure 2A).^27,28^

The *Hybrid brain pulse* likely represents a state in which the arteriole pressure is low but still exceeds the venous pressure. Microvascular blood flow is present but low causing a corresponding fall in pial venule oxygen levels during diastole. The pulse shapes for the 940 nm and 660 nm wavelengths are consequently different due to their distinct responses to low blood oxygen levels (Figure 1B). The Hybrid brain pulse has also been observed in instances of reduced cerebral blood flow both in a study of critically ill brain injury patients during acute falls in blood pressure and in a submitted stroke study in sheep.^14,18^

The *Venous I* and the *Venous II brain pulses* demonstrate venous features and the brain pulse shapes for both 660 and 940 nm are similar. The arteriole and venous pressure levels are likely comparable when Venous I brain pulses are present (Figure 1C). The equilibrium of these two pressure systems results in the Venous I brain pulse having somewhat undifferentiated features which can make the start of the cardiac cycle at times difficult to determine. Venous I may demonstrate more obvious central venous pulse features in the diastolic phase including A and Y waves, as arteriole pressure levels are lower in diastole. The Venous I pulse is consistent with very low cerebral blood flow and perhaps no flow during diastole.

The *Venous II brain pulse* (Figure 1D) has clear central venous features such as the A, C, V, X and Y waves. The Venous II pulse is consistent with venous pressure exceeding arteriole pressure throughout the cardiac cycle which would result in retrograde or reversed blood flow. Reversal of blood flow in cerebral veins has previously been demonstrated in critically ill patients with severe brain injury using transcranial doppler.^29,30^ Reversal of blood flow has also been demonstrated in cortical arterioles, capillary beds and venules in animal models of stroke.^11,31–33^ Flow reversal has also demonstrated in a human cadaver study in response to raised jugular venous pressure.^34^ Pooling of blood in the ipsilateral cerebral veins is commonly present in acute stroke and is associated with worse patient outcomes.^35^ This pooling of venous blood could reflect the low venous pressure levels across the ischemic territory with an inflow of blood from the extensive venous communications within in and between hemispheres.^36^

In a submitted study in a sheep model of stroke we also found Venous class brain pulses were associated with stroke.^18^ In a study of brain injured patients following prolonged out of hospital cardiac arrest we found prolonged Venous II brain pulses were associated with severe brain injury and poor neurological outcome.^14^

The *Monotonous brain pulse* (Figure 1E) has an arterial pressure waveform shape but has some atypical features including a low dicrotic notch and an absence of waveform shape variation over time. Both the low dicrotic notch and the absence of physiological variation over time are consistent with absent cerebral blood flow. Similar characteristics have been demonstrated in intra-cranial pressure waveforms in patients with established brain death.^37,38^ The low dicrotic notch is consistent with absence of a reflected pressure wave from the brain arterioles.^39^ We also found similar features in an ongoing study of brain-dead patients using OBPM.^17^ The mechanism of this pulse may represent a transmitted arterial pressure wave through the brain tissues rather than through the brain’s circulation. The Monotonous brain pulse suggests irreversible brain injury with infarction with absent arterial and venous blood flow. The two patients in our study with Monotonous brain pulses following EVT subsequently had large MCA infarctions on CT imaging and subsequently died. Notably, both cases had extensive bilateral cerebral vascular disease at presentation with poor potential for collateral compensation.

### Assessment of reperfusion and long-term patient outcomes

We found marked differences in the mTICI and the OPBM assessments of reperfusion. Ten of 11 patients obtained a mTICI score of 2B or greater indicating > 50% arterial reperfusion, while only 1 patient demonstrated an improvement in the OBPM brain pulse class following EVT.

The mTICI score is the leading method used to assess arterial reperfusion following EVT but mTICI has limitations in assessing the reperfusion of the microvasculature.^1,6^ These limitations may contribute to why mTICI scores are not consistently predictive of long term patient outcomes.^4^ In our small study, the mTICI scores were not associated with any long-term patient outcomes, while the brain pulse class at the end of EVT was associated with important outcomes, including hospital LOS and mortality, suggesting it could provide additional pathophysiological understanding of the responses to thrombectomy and therefore also assist with neuro-prognostication.

In our cohort, the average time from last seen well to the start of EVT was ∼ five hours, a period expected to trigger microvascular ischemic reperfusion injury, despite successful thrombectomy.^40^ This may result in reduced microvascular blood flow due to leukocyte margination and thrombosis of post-capillary venules with shunting of blood bypassing the thrombosed microvascular beds.^9,11,41^

### Detection of embolization to new territories

One patient in our study had a complication with a large embolization to a new territory during stenting of an internal carotid artery stenosis. The patient demonstrated a clear transition from an Arterial brain pulse to a Venous II brain pulse during stenting. The embolization was detected by routine DSA 9.5 minutes after detected by OBPM. Mechanical thrombectomy successfully removed the emboli 12 minutes later and was associated with improvement of the brain pulse class back to Arterial. This suggests that OBPM could be used to rapidly identify embolization to a new territory between angiography runs, allowing for earlier treatment. The clear improvement in the brain pulse class back to Arterial following the retrieval also suggests that a very short ischemia time may be an important factor in avoiding NRP or sustained ischemic reperfusion injury.

### Contralateral brain pulse changes following LVO stroke

An interesting and perhaps surprising finding was the presence of abnormal brain pulse classes and brain pulse changes on the contralateral hemisphere to the stroke, which were similar to the ipsilateral side. The mechanism of these contralateral changes is unclear but physiological aspects of the brain suggest that pulsatile brain movement could be one factor. The brain is one of the softest tissues in the human body and deforms under its own weight and floats in cerebrospinal fluid.^42^ Cerebral blood flow, central venous pressure, cerebrospinal fluid flow and breathing can all induce brain motion in this delicate organ.^43,44^ In cases of brain injury, distinct and abnormal brain motions are observed across both hemispheres associated with the cardiac pulse and breathing.^44,45^ We speculate that the contralateral changes may represent pulsatile brain movement induced by a large portion of the ipsilateral blood volume under the influence of venous pressure. The venous circulation represents 70% of the entire brain blood volume and there are extensive venous communications between hemispheres.^36^ Other mechanisms could also contribute including steal of arterial blood flow from the contralateral side.^46^ Human and animal studies have demonstrated reductions of cerebral blood flow in the contralateral hemisphere. In a human study cerebral blood flow fell by 35% at 8 hours following stroke. and ∼ 20% in an animal model.^47,48^ In this study, CT perfusion only demonstrated impaired perfusion on the contralateral side in one patient, though this imaging technique represents a relative difference in perfusion rather than absolute level of perfusion, meaning it does not exclude the possibility of a reduction contralateral perfusion in the other patients.

### LVO Stroke detection

We found that waveforms with prominent venous pressure features were present in acute LVO stroke which suggests OBPM could assist in earlier diagnosis of stroke allowing for faster and improved clinical decision making in these resource limited settings. Limited access to brain imaging is unfortunately common in low population, less well-resourced regions resulting in delayed stroke detection and treatment and consequently worse long-term patient outcomes.^49^ A simple, inexpensive point of care monitor, in conjunction with telemedicine stroke specialists, could assist in an earlier decision for transport to a comprehensive stroke unit and treatment could occur earlier with potential to improve patient outcomes.

### Limitations

A major limitation was the small size of this study; therefore, the analyses were exploratory in nature and a larger study is required to confirm the current findings. The study also covered a short window of time course of EVT. Monitoring the brain responses for longer periods could have provide better understanding of the pathophysiological responses following EVT, which could also include the detection of post-procedural complications, which occur in up to 25% of cases.^50^

In this study, the class of the brain pulse was classified by blinded investigators. The brain pulse waveforms from the OBPM reflect a continuum based on arterial and venous pressure in the brain and consequently there can be subjectivity when dividing into classes. Misclassifications are possible particularly with arterial and monotonous which can have similar features. Objective signal processing approaches should be considered in a future study.

No patients had formal assessment of NRP using CT perfusion or MR perfusion, which could have provided additional understanding of the role of OBPM in detecting NRP. Though currently no gold standard approach exists to assessing the microcirculation and no reflow phenomenon.^5^

The sensors were only placed over the MCA territories which provide the best signal as the bone is thin in the temporal area and future studies could assess other territories.

## Conclusion

This first-in-human study found the OBPM brain pulse has venous circulation features in acute LVO stroke. During EVT the brain pulse waveforms provided insights into the microcirculation and blood flow changes associated with EVT. These waveforms were associated with important clinical outcomes, such as stroke size at presentation, hospital LOS and mortality. Taken together, the waveforms may allow for earlier detection of complications during EVT compared to standard DSA, provide better assessment of NRP and improved neuro-prognostication. This real-time monitoring during EVT therefore could enhance stroke management, enabling clinicians to make earlier decisions and improve patient outcomes. Finally, our findings suggest OBPM may provide a simple point of care method for LVO stroke detection in resource limited settings.

## Data Availability

All data produced in the present study are available upon reasonable request to the authors

## Acknowledgements

We would like to thank the staff of St Vincent’s Hospital Melbourne for supporting us to undertake this research.

## Supplemental Material

Tables S1-S3

Figures S1-S3

## Non-standard Abbreviations and Acronyms

OBPM: optical brain pulse monitor(ing)
EVT: endovascular treatment
LVO: large vessel occlusion
mTICI: modified treatment in cerebral ischemia
LOS: length of stay
MCA: middle cerebral artery
ICA: internal carotid artery
DSA: digital subtraction angiography
CT: computed tomography
MRI: magnetic resonance imaging
NRP: no reflow phenomena
NIHSS: NIH stroke scale
mRS: modified Rankin Scale
HREC: human research ethics committee
GUI: Graphical User Interface

## Graphical Abstract

Optical brain pulse waveforms were obtained in 11 stroke patients undergoing EVT. At the start of EVT, the ipsilateral brain pulse class was associated with the hypoperfused tissue volume. In one patient, an abrupt transition from Arterial to Venous brain pulse during carotid stenting indicated emboli to new territory, detectable before standard DSA. Ipsilateral brain pulse class following EVT was associated with longer LOS and mortality. *Abbreviations: EVT, endovascular therapy; DSA, digital subtraction angiography; CT, computed tomography*.

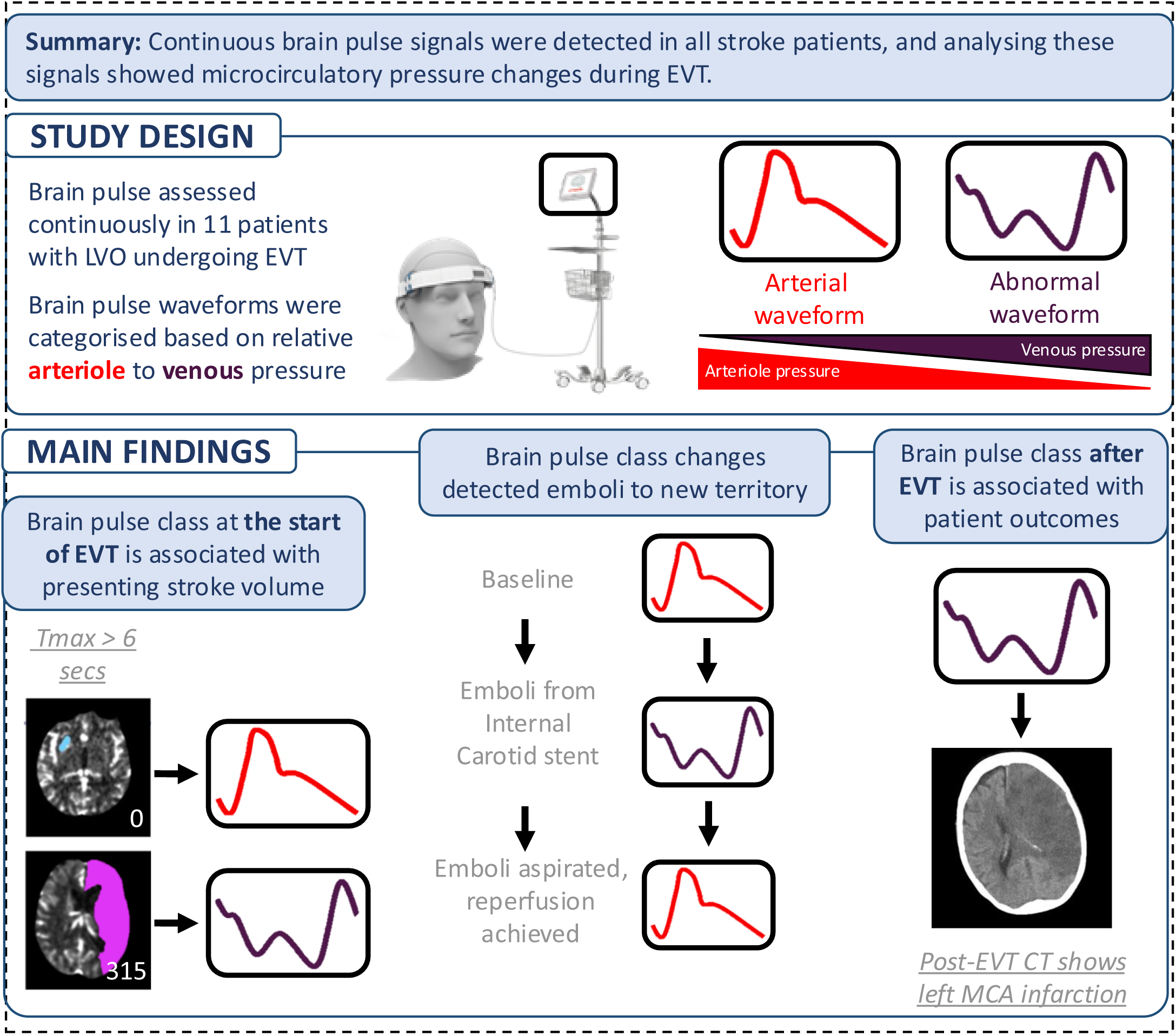

