## Supplementary material for "Optical brain pulse monitoring of microvascular blood flow during endovascular treatment for acute ischemic stroke": Supp Tables 1-3 and Supp Figures 1-3

**Supplementary Table 1. Summary of patient outcomes**

| **Patient** | **Episode*** | **mTICI** | **No. passes** | **LOS (days)** | **Hospital death** | **90-day mRS** |
| --- | --- | --- | --- | --- | --- | --- |
| 1 | 1 | 3 | 2 | 25 | No | 3 |
| 1 | 2 | 3 | 2 |  |  |  |
| 2 | 1 | 2B | 3 | 22 | No | 3 |
| 3 | 1 | 3 | 1 | 3 | No | 2 |
| 4 | 1 | 3 | 1 | 55 | No | 4 |
| 5 | 1 | 2B | 1 | 3 | Yes | 6 |
| 6 | 1 | 1 | 4 | 17 | Yes | 6 |
| 7 | 1 | 2C | 4 | 3 | Yes | 6 |
| 8 | 1 | 2C | 4 | 28 | No | 1 |
| 9 | 1 | 3 | 3 | 11 | No | 2 |
| 10 | 1 | 3 | 3 | 10 | No | 2 |
| 11 | 1 | 2B | 1 | 18 | No | 3 |

* Episode is defined as the number of EVT procedures a patient underwent during their hospital stay. For example, episode 1 refers to the patient’s first EVT, while episode 2 refers to their second EVT.

*Abbreviations: mTICI, modified treatment in cerebral infarction; LOS, length of stay; mRS, modified Rankin Scale; EVT, endovascular treatment.*

**Supplementary Table 2. Endovascular treatment procedural data**

| **Patient** | **Episode*** | **tPA** | **Intubated** | **Time from last seen well to start of EVT** |
| --- | --- | --- | --- | --- |
| 1 | 1 | No | No | 3 hrs 21 mins |
| 1 | 2 | No | No | 1 hr 42 mins |
| 2 | 1 | No | Yes | 1 hr 30 mins |
| 3 | 1 | No | Yes | 18 hrs |
| 4 | 1 | No | Yes | 43 mins |
| 5 | 1 | Yes | Yes | 3 hrs 35 mins |
| 6 | 1 | No | Yes | 17 hrs |
| 7 | 1 | No | Yes | 1 hr 25 mins |
| 8 | 1 | Yes | Yes | 1 hr 45 mins |
| 9 | 1 | No | Yes | 1 hr 27 mins |
| 10 | 1 | No | Yes | 9 hrs 24 mins |
| 11 | 1 | No | Yes | 1 hr 30 mins |

* Episode is defined as the number of EVT procedures a patient underwent during their hospital stay. For example, episode 1 refers to the patient’s first EVT, while episode 2 refers to their second EVT.

*Abbreviations: tPA, tissue plasminogen activator; EVT, endovascular treatment.*

**Supplementary Table 3. Contralateral brain pulse waveform classifications over the course of EVT.**

| *Patient* | *Episode** | *Start of EVT* | *During clot retrieval* | *Post-clot  retrieval* | *End of EVT* | *Change in classification* |
| --- | --- | --- | --- | --- | --- | --- |
| 1 | 1 | Venous I | Venous I | Venous I | Venous I | No change |
| 1 | 2 | Venous I | Arterial | Arterial | Venous I | Reversion |
| 2 | 1 | Venous I | Venous II | Venous II | Venous II | Worsened |
| 3 | 1 | Arterial | Arterial | Venous I | Arterial | Reversion |
| 4 | 1 | Venous I | Venous I | Venous I | Venous I | No change |
| 5 | 1 | Venous I | Venous I | Venous I | Venous I | No change |
| 6 | 1 | Venous I | Venous I | Venous I | Venous I | No change |
| 7 | 1 | Venous I | Venous I | Venous II | Venous II | Worsened |
| 8 | 1 | Arterial | Arterial | Arterial | Arterial | No change |
| 9 | 1 | Venous I | Venous I | Venous I | Venous I | No change |
| 10 | 1 | Arterial | Venous I | Venous II | Arterial | Reversion |
| 11 | 1 | Venous I | Venous I | Hybrid | Venous I | Reversion |

* Episode is defined as the number of EVT procedures a patient underwent during their hospital stay. For example, episode 1 refers to the patient’s first EVT, while episode 2 refers to their second EVT.

*Abbreviations: tPA, tissue plasminogen activator; EVT, endovascular treatment.*

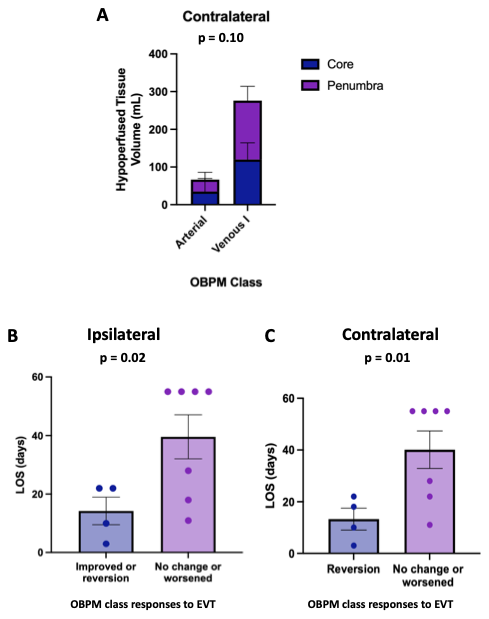

**Supplementary Figure 1. OBPM Class at presentation and in responses to EVT:** (A) Hypoperfused stroke volume was not associated with contralateral brain pulse waveform class at presentation (p = 0.10). The components of the hypoperfused stroke volume (Tmax > 6 seconds), Core (<30% normal cerebral blood flow) and penumbra are demonstrated. (B & C) Patients that demonstrated either an improvement in the OBPM pulse class or a brief period of deterioration followed by reversion to the original brain pulse class during EVT had a shorter hospital LOS compared to patients in which the class did not change or worsened during EVT. (B) Ipsilateral brain pulse class response (p=0.02). (C) Contralateral brain pulse class response (p=0.01). Assessed using Welch’s t-test. *Abbreviations: LOS, length of stay. EVT, endovascular therapy; OBPM, optical brain pulse monitor.*

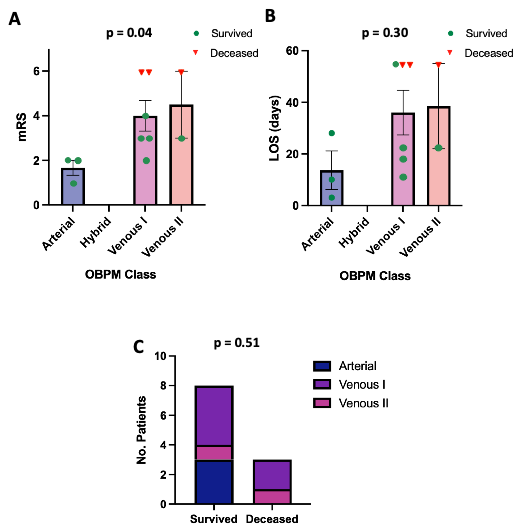

**Supplementary Figure 2. Long-term Patient Outcomes associations with Contralateral Optical Brain Pulse Class at end of EVT.** (A) The 90-day mRS score (p = 0.04), (B) adjusted LOS in days (p = 0.30), and (C) In-hospital mortality (p = 0.51). OBPM demonstrated statistical association with mRS only, assessed using Welch’s t-test (A & B) and (C) Fisher’s exact test. *Abbreviations: LOS, length of stay; EVT, endovascular therapy; mRS, modified Rankin Scale.*

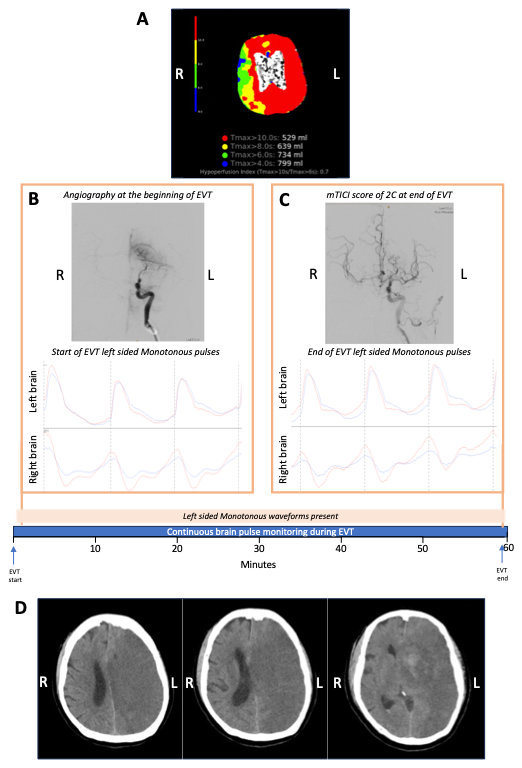

**Supplementary Figure 3:** **Monotonous waveforms observed during EVT in a patient with poor neurological outcome**. (A) Rapid CT perfusion imaging at presentation reveals a large hypoperfused volume. (B) Angiography at presentation shows an occlusion of the left ICA. OBPM recordings indicate left-sided Monotonous and right-sided Venous I brain pulses. (C) Post-EVT, DSA demonstrates effective arterial recanalization with mTICI 2C achieved; however, OBPM shows no change in waveform. Faint grey dashed lines mark the onset of each brain pulse. (D) Follow-up CT imaging 2 days post-EVT reveals infarct of the entire left hemisphere with midline shift. The patient did not survive. *Abbreviations: DSA, digital subtraction angiography; EVT, endovascular therapy; mTICI, modified treatment in cerebral infarction; CT, computed tomography; ICA, internal carotid artery; L, left; R, right. Red pulse: 660 nm; Blue pulse: 940 nm.*
